# Proteome-Wide Assessment of Clustering of Missense Variants in Neurodevelopmental Disorders Versus Cancer

**DOI:** 10.1101/2024.02.02.24302238

**Authors:** Jeffrey K. Ng, Yilin Chen, Titilope M. Akinwe, Hillary B. Heins, Elvisa Mehinovic, Yoonhoo Chang, Zachary L. Payne, Juana G. Manuel, Rachel Karchin, Tychele N. Turner

**Affiliations:** Department of Genetics, Washington University School of Medicine, St. Louis, MO 63110, USA; Department of Biomedical Engineering, Johns Hopkins University, Baltimore, MD, USA; Molecular Genetics & Genomics Graduate Program, Washington University School of Medicine, St. Louis, MO 63110, USA; Human & Statistical Genetics Graduate Program, Washington University School of Medicine, St. Louis, MO 63110, USA; Department of Computer Science, Johns Hopkins University, Baltimore, MD, USA; The Sidney Kimmel Comprehensive Cancer Center, School of Medicine, Johns Hopkin University, Baltimore, MD, USA; Institute for Computational Medicine, Johns Hopkins University, Baltimore, MD, USA; Intellectual and Developmental Disabilities Research Center, Washington University School of Medicine, St. Louis, MO, USA

**Author notes:** co-first authors (Jeffrey K. Ng, Yilin Chen, Titilope M. Akinwe). Correspondence to &; Tychele N. Turner, Ph.D., Washington University School of Medicine Department of Genetics, 4523 Clayton Avenue, Campus Box 8232 St. Louis, MO 63110; Rachel Karchin, Ph.D. Johns Hopkins University, Departments of Biomedical Engineering and Computer Science Institute for Computational Medicine, Baltimore, MD 21205.

## Abstract

Missense de novo variants (DNVs) and missense somatic variants contribute to neurodevelopmental disorders (NDDs) and cancer, respectively. Proteins with statistical enrichment based on analyses of these variants exhibit convergence in the differing NDD and cancer phenotypes. Herein, the question of why some of the same proteins are identified in both phenotypes is examined through investigation of clustering of missense variation at the protein level. Our hypothesis is that missense variation is present in different protein locations in the two phenotypes leading to the distinct phenotypic outcomes. We tested this hypothesis in 1D protein space using our software CLUMP. Furthermore, we newly developed 3D-CLUMP that uses 3D protein structures to spatially test clustering of missense variation for proteome-wide significance. We examined missense DNVs in 39,883 parent-child sequenced trios with NDDs and missense somatic variants from 10,543 sequenced tumors covering five TCGA cancer types and two COSMIC pan-cancer aggregates of tissue types. There were 57 proteins with proteome-wide significant missense variation clustering in NDDs when compared to cancers and 79 proteins with proteome-wide significant missense clustering in cancers compared to NDDs. While our main objective was to identify differences in patterns of missense variation, we also identified a novel NDD protein BLTP2. Overall, our study is innovative, provides new insights into differential missense variation in NDDs and cancer at the protein-level, and contributes necessary information toward building a framework for thinking about prognostic and therapeutic aspects of these proteins.

## INTRODUCTION

Neurodevelopmental disorders (NDDs) affect ~1% of the population and include autism, developmental delay, epilepsy, and intellectual disability. There have been several studies that have identified genetic contributions to NDDs including common variants (1), rare inherited variants (2, 3), and *de novo* variants (DNVs) (4–43). In particular, the study of DNVs through enrichment at the gene-level (44) has identified >300 genes involved in NDDs (36, 43). The initial discoveries were genes with high-impact likely gene-disruptive (LGD) DNVs (e.g., stop-gain, splice-site acceptor, splice-site donor, frameshift) (22, 45). The estimation of gene discovery from DNVs suggests a plateau at ~30,000 parent-child sequenced trios for LGD DNVs and for bioinformatically predicted severe missense DNVs a plateau at ~10,000 parent-child sequenced trios. However, where the plateau will be for gene discovery for the broader class of missense DNVs contributing to NDDs has been difficult to determine as they are harder to interpret than LGD DNVs (36). This highlights the importance for the development of computational and statistical tools to study this class of variation. Identification of relevant missense DNVs is critical for prognostics, functional testing, and future therapeutic strategies.

It is generally accepted that recurrent missense variants at a single amino acid residue position or in close proximity in a protein is a signature of strong positive selection (46). This hypothesis has motivated the search for clusters of driver mutations in cancers, both in one-dimensional gene or protein sequence and three-dimensional protein structures (47–50). While clusters of driver mutations have primarily been associated with oncogenes (51), we have previously shown that these clusters occur in both oncogenes and tumor suppressor genes (50). Furthermore, we developed the software CLUMP to compare clustering of germline variants in autosomal dominant vs. recessive Mendelian diseases and we identified more clustering of missense variants in autosomal dominant than autosomal recessive diseases (52). Application of CLUMP to NDDs has identified proteins with clustering of missense DNVs (53), and other groups have utilized other strategies to assess clustering of missense DNVs in NDDs at the gene or protein level (43, 54, 55).

In this study, we 1) apply the original CLUMP method to perform case-control testing to examine clustering in 1D protein space (**Figure 1A**), 2) and develop a new method called 3D-CLUMP to perform case-control testing to examine clustering in three-dimensional (3D) protein structure (**Figure 1B**). Both strategies are advantageous because they do not use *a priori* information about domains or known regions of functional importance within a protein.

**Figure 1:**
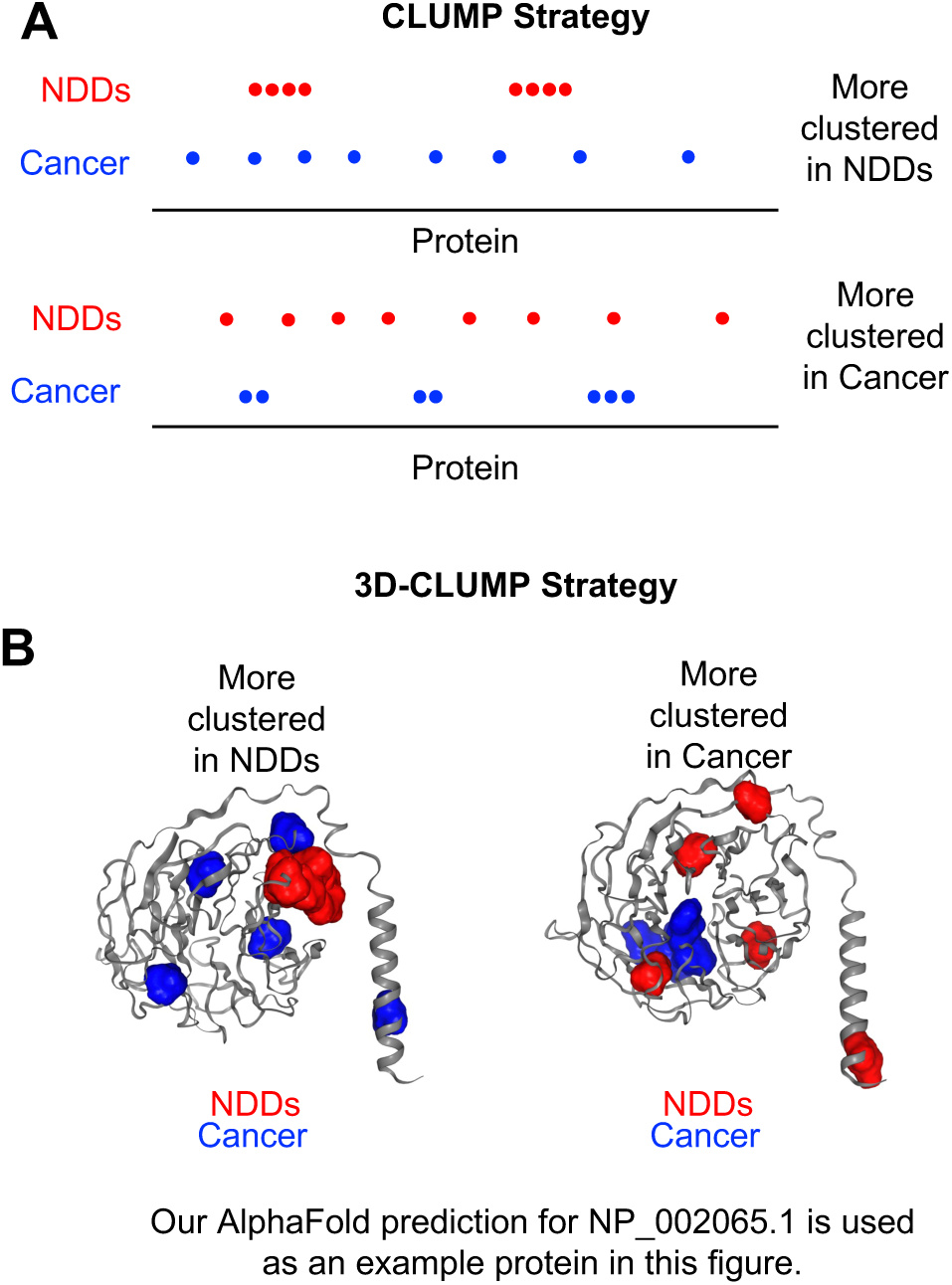
Schematic of Examples of the CLUMP and 3D-CLUMP Methods. A) Two proteins are shown: one where there is more clustering in NDDs (Top) and one where there is more clustering in cancer (Bottom). B) Our AlphaFold prediction for NP_002065.1 (used as an example only) is shown in this image where variants are placed to exemplify more clustering in NDDs (Left) and more clustering in cancer (Right).

As syndromic and genome-wide significant NDD genes have been identified, there has been an interesting observation that some of these genes are also implicated in cancers. Initial examples included *PTEN* in Cowden syndrome and *NF1* in Neurofibromatosis I (56, 57). More recent examples include *CHD8* in autism and in gastrointestinal cancers (45). However, for the majority of genes it remains unclear whether the specific variants identified in NDDs will later lead to cancer or not. In 2016, a forum paper by Crawley et al. (57) described an overlap of genes involved in both autism and cancer. They noted the centering of these genes on molecular pathways involved in gene regulation (e.g., chromatin, transcription, signaling). This was further examined in a review paper by Nussinov et al. in 2022 (58). Understanding why some of the same genes are involved in both NDDs and cancer is essential for prognostics and future therapeutics. In this study, we test the hypothesis that some genes involved in both NDDs, and cancer exhibit differential missense DNV clustering in the two phenotypes. To test this hypothesis, we aggregated NDD DNVs from the literature (37, 43) and cancer somatic variants from The Cancer Genome Atlas (TCGA) and Catalog Of Somatic Mutations in Cancer (COSMIC) databases and tested them using our existing (i.e., CLUMP (52) and our newly developed (3D-CLUMP) clustering tools.

In this study, our objectives are to identify 1) proteins which exhibit missense clustering in NDDs and not cancer and 2) proteins that exhibit missense clustering in cancer and not NDDs. This work involves the development of a new computational tool of broader use to the research community for assessing clustering of missense variation in 3D protein structures and provides novel insights into variation involved in cancer and NDDs with potential prognostic and therapeutic implications.

## MATERIALS AND METHODS

### Annotation of Variant Data

We aggregated DNV data from 39,883 parent-child sequenced NDD trios (37, 43) (**Figure 2A**). The DNVs were annotated with VEP (59) to RefSeq protein isoforms. Somatic variants (**Figure 2B**) were downloaded from TCGA at https://gdc.cancer.gov/about-data/publications/mc3-2017 for the following cancer types: Breast Cancer (BRCA), Colon Adenocarcinoma (COAD), Lung Adenocarcinoma (LUAD), PRAD (Prostate Adenocarcinoma), and Ovarian Cancer (OV). Somatic variants were also downloaded from the COSMIC database at https://cancer.sanger.ac.uk/cosmic/download in which multiple tissue types were aggregated in the categories of CNS (Central Nervous System) or GI (Gastrointestinal Track). CNS tissue types: Aqueduct of Sylvius, Basal ganglia, Brain Brainstem, Caudate nucleus, Cerebral cortex, Cerebral hemisphere, Cerebrum Chiasm, Choroid plexus, Cingulate gyrus, Cingulum, Corpus callosum, Diencephalon, Extra-central nervous system, Filum Foramen of Monro, Fourth ventricle, Frontal lobe, Frontobasal, Frontoparietal, Frontotemporal, Hypothalamus, Infratentorial, Intraventricular, Lateral ventricle, Lateral ventricle trigone, Left hemisphere, Medulla, Medullo cerebellar, Meninges, Midbrain, Middle cerebellar peduncle, Middle frontal gyrus, NS, Occipital lobe, Optic nerve, Optic pathway, Paracentral Parietal lobe, Parietooccipital, Parietotemporal, Periventricular, Pineal gland, Posterior fossa, Precentral gyrus, Sella turcica, Sellar suprasellar, Septum pellucidum, Spinal cord, Subcorticoparamedial, Superior frontal gyrus, Supratentorial, Tectum, Tegmentum, Temporal lobe, Temporobasal, Temporooccipital, Temporoparietal, Thalamus, Third ventricle, Trigone. GI tissue types: Large_intestine, Small_intestine, Gastrointestinal track undetermined site, Esophagus, and Stomach. The somatic variants were also annotated with VEP to RefSeq protein isoforms.

**Figure 2:**
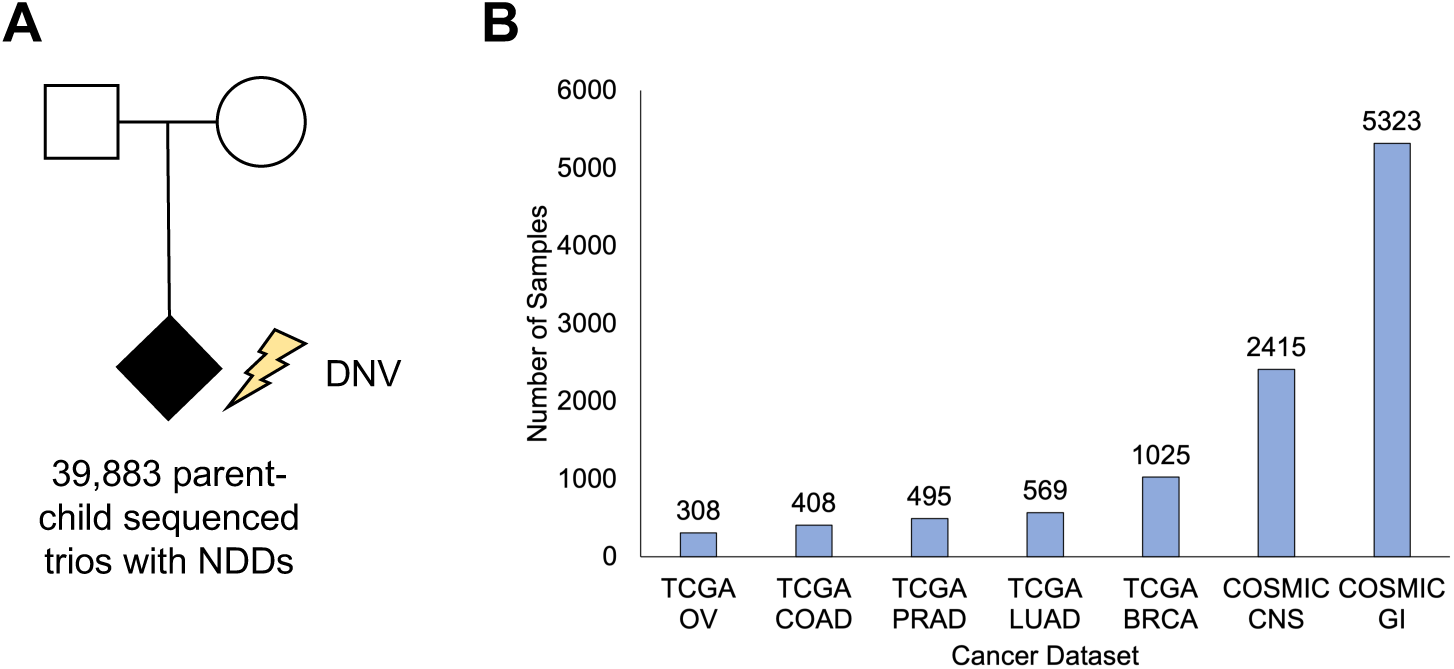
Variant Data Types in This Study. A) NDD data consisted of 39,883 parent-child sequenced trios (the lightning bolt is used to exemplify DNVs which, by definition, are only found in children). B) Cancer data consisted of 10,543 individuals from the TCGA and COSMIC databases.

### Application of Case-Control CLUMP

To assess clustering in 1D protein space, CLUMP was utilized for comparison of NDDs and cancer. CLUMP was run to compare missense variation identified in individuals with NDDs to missense variants identified in each cancer type, respectively. For this analysis, CLUMP was run in the case-control implementation using a *-m* value of 6 to denote >5 missense variants required in each dataset and a *-z* value of 10,000,000 to signify a permutation of 10 million for significance testing. This permutation level allows for proteome-wide significance testing in the dataset.

### AlphaFold Structure Prediction for Proteins with Missense Variation

We downloaded the RefSeq protein fasta file (https://ftp.ncbi.nlm.nih.gov/refseq/H_sapiens/annotation/GRCh38_latest/refseq_identifiers/GRC <u>h38_latest_protein.faa.gz</u>) from NCBI on May 18, 2021. For each protein, we extracted the protein sequence in fasta format from the RefSeq file. To generate a 3D protein structure for the protein, the protein fasta file was used as input to the AlphaFold (60) version 2.2.0 program. The following reference databases were used: *Reduced BFD*, *MGnify (2018_12)*, *Uniclust30 (2018_08)*, *UniRef90 (2022_01)*, and *PDB70 (2020-04-01)*. The AlphaFold program generates an MSA using the *reduced_dbs* setting, model generation produces 5 models with 3 recycles each and AMBER relaxation. The models are ranked by pLDDT, and the best scoring structure is chosen as the final structure. Post-AlphaFold structure generation, each structure was deposited under accession “matur-clump” in ModelArchive. Proteins requiring >700 GB of memory were unable to be run through AlphaFold on our server.

### The 3D-CLUMP Method

Development of 3D-CLUMP as a strategy to assess clustering of variation in 3D protein structures required modification of the method we originally developed for 1D clustering called CLUMP (52). As in the CLUMP method, 3D-CLUMP uses a partitioning around medoids (PAM) strategy specifically using the *pamk* function from the R package fpc (https://cran.r-project.org/web/packages/fpc/index.html) to identify clusters in the data. The advantage of integrating *pamk* in 3D-CLUMP is that it iteratively identifies the optimal number of clusters *k* in the data to find the optimal *k\**. This is helpful as the users of the program may not know the optimal *k* for their dataset. The *pamk* function identifies both the number of clusters and estimates the ‘medoid’ that best represents each cluster. Importantly, in 3D-CLUMP the medoid is represented in 3D-space consisting of x,y,z coordinates on the 3D protein structure. The final 3D-CLUMP score (*Sp*) for a protein *p* is calculated as follows:

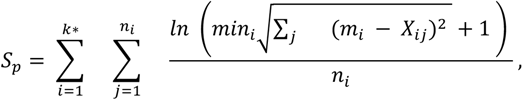

where *k\** is the optimal *k* for the protein, *n_i_* is the number of variants in cluster *i*, *m_i_* is the position of the medoid in the *i* cluster, and X_ij_ is the position of variant *j* in the *i* cluster. If all variants cluster at the exact same location as the medoid, this will yield optimal clustering and will result in a score of 0. In 3D-CLUMP, lower scores show maximal clustering of the variants.

To calculate a p-value for the test, we generate a null distribution of 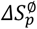 values, which is 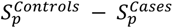. The 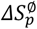 values are calculated 10,000,000 times to enable testing for proteome-wide (i.e., genome-wide correction for 19,008 genes) significance. We use a Bonferroni corrected 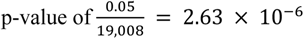 as the threshold for proteome-wide significance.

### Application of Case-Control 3D-CLUMP

To assess clustering in 3D protein space, 3D-CLUMP was utilized for comparison of NDDs and cancer. 3D-CLUMP was run to compare missense variation identified in individuals with NDDs to missense variants identified in each cancer type, respectively. For this analysis, 3D-CLUMP was run in the case-control implementation using a *-m* value of 6 to denote >5 missense variants required in each dataset and a *-z* value of 10,000,000 to signify a permutation of 10 million for significance testing. This permutation level allows for proteome-wide significance testing in the dataset.

### Plotting of Variant Data

Missense variants were visualized on 3D structures using the R package *NGLVieweR* (https://cran.r-project.org/web/packages/NGLVieweR/vignettes/NGLVieweR.html). Missense variants identified in individuals with NDDs were colored red in the plots and missense variants identified in individuals with cancer were colored blue in the plots. The intensity of the color corresponds to the number of individuals with DNVs at the specific amino acid residue.

## RESULTS

### Proteins Exhibiting Clustering in 1D Protein Space

For each of the seven cancer types, testing for significance of clustering in 1D space was done with CLUMP. For the comparison to BRCA, we identified two proteins reaching significance in NDDs (ADCY5, SETD2) and one protein reaching significance in cancer (PIK3CA). For the comparison to COAD, we identified four proteins reaching significance in NDDs (ALG13, CREBBP, PACS1, UPF1) and no protein reaching significance in cancer. For the comparison to LUAD, we Identified 9 proteins reaching significance in NDDs (ADCY5, ALG13, CREBBP, EBF3, FOXP1, ITPR1, PPP2R1A, PPP2R5D, TRIO) and no protein reaching significance in cancer. For the comparison to PRAD, we identified one protein reaching significance in NDDs (DYNC1H1) and no protein reaching significance in cancer. For the comparison to OV, we did not identify any proteins reaching significance in either NDDs or cancer. For the comparison to COSMIC CNS, we identified one protein reaching significance in NDDs (PPP2R5D) and 17 protein reaching significance in cancer (ATP8A1, BAHCC1, DMD, FAM186A, IGSF10, KALRN, KANK1, KDM3B, MADD, MUC6, NEB, NWD2, PLXNB2, SRRM2, TBCEL-TECTA, TRPM7, UNC80). For the comparison to COSMIC GI, we identified six proteins reaching significance in NDDs (ARID1B, COG4, DYNC1H1, PTPN11, TRIO, TRPM3) and five protein reaching significance in cancer (GNAS, KALRN, MUC19, PIK3CA, SPTA1).

### Metrics of AlphaFold Structures

Summary metrics of the AlphaFold structures generated for the dataset include a pLDDT of 69.79 ± 21.56 (mean ± standard deviation). Across the dataset, there was a negative correlation between the mean pLDDT value and the standard deviation pLDDT values (r = −0.33, p < 2.2 × 10^−16^), a negative correlation between the protein length and the mean pLDDT values (r = −0.31, p < 2.2 × 10^−16^), and a positive correlation between the protein length and the standard deviation pLDDT values (r = 0.21, p < 2.2 × 10^−16^). While the correlations were significant they were not strong. Observation of the data revealed there was an apparent inverse U-shape to the data when comparing the standard deviation pLDDT to the mean pLDDT. To test this we fit the standard deviation and mean to a model using the following formula in R:

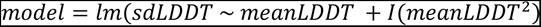

This model fit the data well with an adjusted *r*^2^ = 0.61 and a p < 2.2 × 10^−16^. This result shows that structures with good predictions overall (i.e., high mean pLDDT scores) tend to have lower standard deviation for their pLDDT scores and structures with bad predictions overall (i.e., low mean pLDDT scores) tend to have lower standard deviation for their pLDDT scores. In short, really good structures are good overall and really bad structures are bad overall. There are then several structures with decent structures but lots of variability in how consistent they are in pLDDT overall.

### Proteins Exhibiting Clustering in 3D Protein Space

For each of the seven cancer types, testing for significance of clustering in 3D space was done with 3D-CLUMP (**Figure 3**, **Figure 4**). For the comparison to BRCA, we identified 15 proteins reaching significance in NDDs (ALG13, ATP1A3, BLTP2, BRAF, CACNA1A, CDK13, DDX3X, HDAC4, MAPK8IP3, PACS1, SCN3A, SCN8A, SETBP1, SMARCA2, TCF4) and one protein reaching significance in cancer (PIK3CA). For the comparison to COAD, we identified 23 proteins reaching significance in NDDs (ACTL6B, ALG13, ATP1A3, ATP6V0A1, BLTP2, COG4, CSNK2A1, DDX23, DHX30, DYRK1A, GRIN1, HECW2, KCNK3, KCNQ3, KCNT2, KIF1A, MECP2, MEF2C, PACS1, PTPN11, SCN2A, SMARCA2, TRAF7) and no protein reaching significance in cancer. For the comparison to LUAD, we identified 22 proteins reaching significance in NDDs (ACTL6B, ALG13, ATP6V0A1, BLTP2, FBXW7, FGFR2, FOXP1, KCNQ3, KCNT2, KIF1A, MECP2, MEF2C, PPP2R1A, PPP2R5D, PTPN11, SCN2A, SCN8A, SMAD4, SMARCA2, TCF4, TFE3, TRIP12) and one protein reaching significance in cancer (KIDINS220). For the comparison to PRAD, we identified two proteins reaching significance in NDDs (KIF1A, SMAD4) and one protein reaching significance in cancer (SPOP). For the comparison to OV, we identified three proteins reaching significance in NDDs (BLTP2, SCN2A, SCN3A) and no protein reaching significance in cancer. For the comparison to COSMIC CNS, we identified six proteins reaching significance in NDDs (ALG13, CDK13, FBXW7, HDAC4, PPP2R5D, SMARCA2) and 54 proteins reaching significance in cancer (ABCA3, ABCA7, AGAP1, AGAP2, ANK2, ATP1A2, ATP8A1, CABIN1, CACNA1B, CIC, CLASP1, COQ8A, CTBP2, DICER1, DIDO1, DOCK7, EPHB4, ESYT1, FAM83H, FLII, HELZ, KANK1, KIDINS220, KIF19, KMT2E, LAMB1, MADD, MAST2, MPDZ, MYO16, NCOR1, NWD2, PCDH15, PCNX3, PHRF1, PIK3CA, PKDREJ, PLXNB2, PTPRS, PTPRZ1, RALGAPA1, SCN1A, SCRIB, SLX4, SNRNP200, STAT1, TCF20, TRIM41, TRIP12, TRPM7, USH2A, VCP, ZC3H13, ZSWIM6). For the comparison to COSMIC GI, we identified 18 proteins reaching significance in NDDs (ALG13, ATP1A3, BLTP2, CDK13, DHDDS, DHX30, FGFR3, HDAC4, HK1, KCNK3, KCNT1, KCNT2, KIF1A, MAPK8IP3, NAA10, PTPN11, SMARCA2, SOS1) and 13 protein reaching significance in cancer (ABCA8, ABHD16A, CSPP1, GNAS, KANK1, NLRP13, PIK3CA, PTEN, TP73, TTLL10, ZFYVE28, ZHX3, ZNF438).

**Figure 3:**
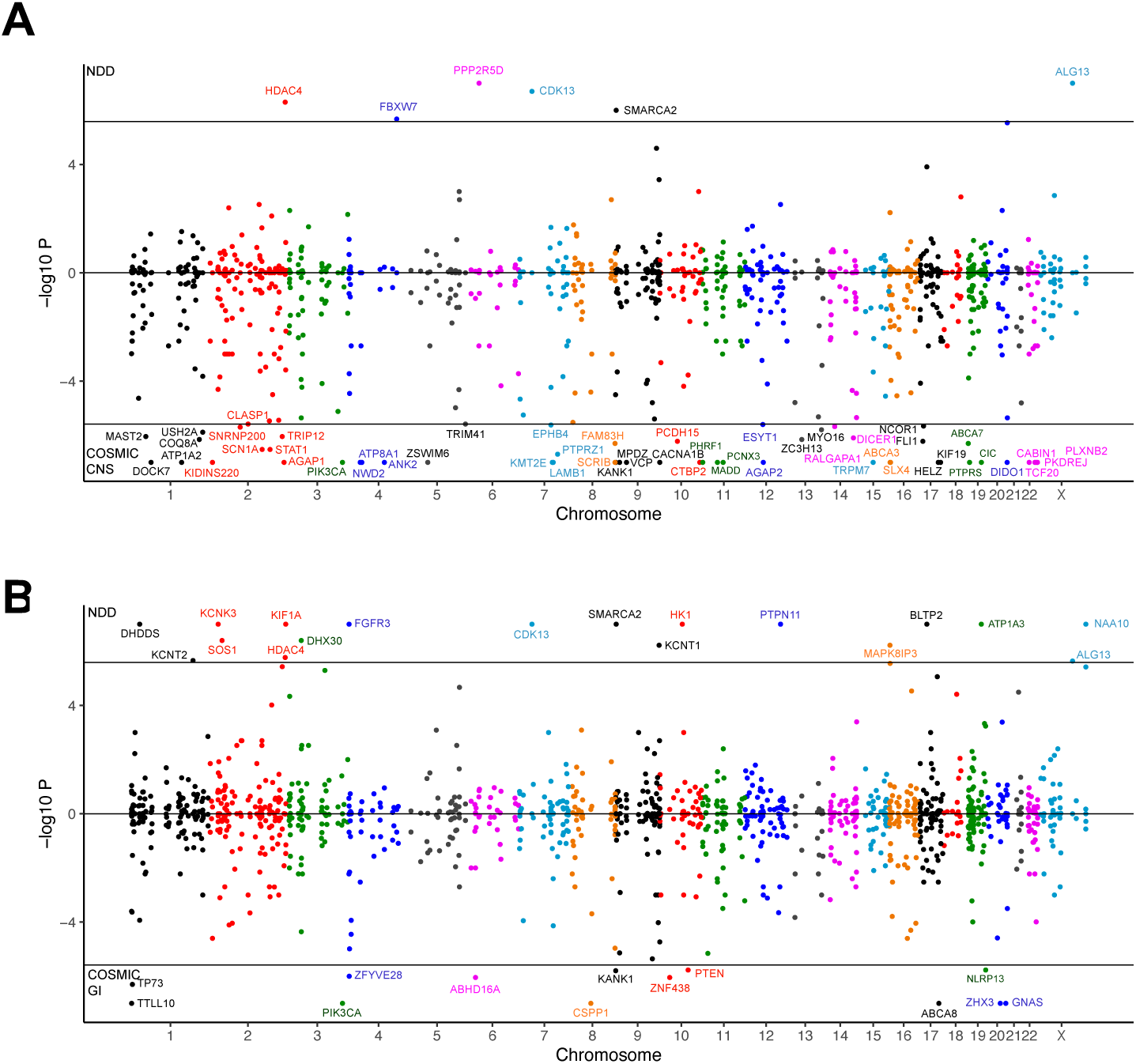
Chicago Plots for the 3D-CLUMP Results in the COSMIC Datasets. A) Chicago plot for 3D-CLUMP results in the NDD versus COSMIC CNS analyses. B) Chicago plot for 3D-CLUMP results in the NDD versus COSMIC GI analyses. For both A and B proteins that exhibit significant clustering in NDDs are shown on the top above the significance line and proteins that exhibit significant clustering in cancer are shown on the bottom below the significance line. Proteins are placed based on the genomic coordinates of the genes that encode them and all significant proteins are labeled on the plots.

**Figure 4:**
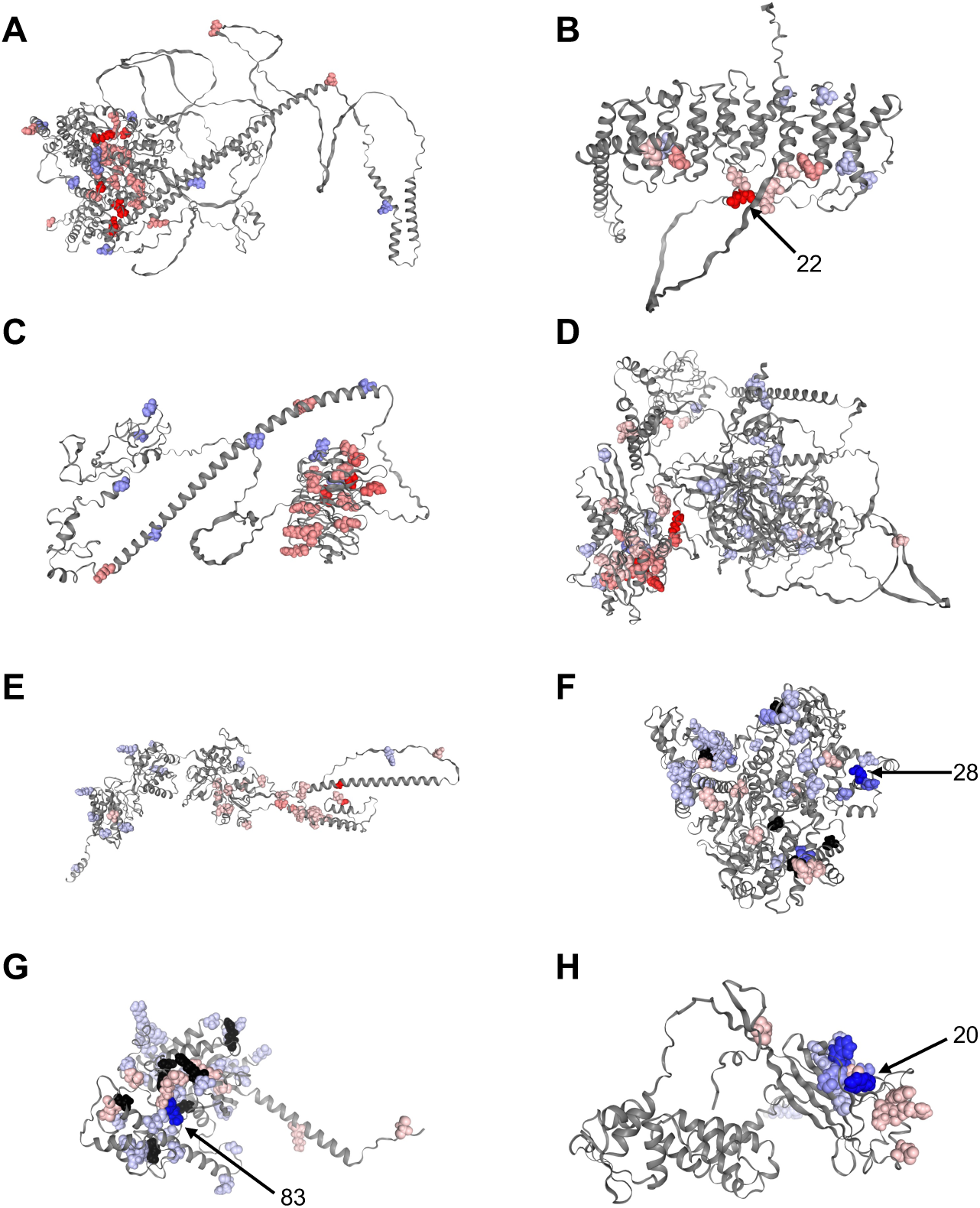
Examples of Proteins with Proteome-Wide Significant Clustering of Missense Variants. Subfigures A to E are significant in NDDs and subfigures F to H are significant in cancer. Red are variants seen in individuals with NDDs. Numbers are shown next to some residues to indicate the number of individuals. Blue are seen in individuals with cancer. Black are seen in both. The intensity of the color is scaled by the number of individuals with missense variants at the residue. A) NDD versus BRCA SMARCA2 (NP_620614), B) NDD versus CNS PPP2R5D (NP_851307), C) NDD versus COAD TRAF7 (NP_115647), D) NDD versus LUAD KIF1A (NP_004312), E) NDD versus COAD GRIN1 (NP_067544), F) NDD versus CNS PIK3CA (NP_006209), G) NDD versus GI GNAS (NP_000507), H) NDD versus PRAD SPOP (NP_003554).

### Greater Statistical Discovery Through Assessment of Clustering in 3D-Structures

Using the 3D protein structure test (3D-CLUMP), greater significant protein discovery was observed in comparison to the 1D test. In every comparison of NDDs and the shown cancer type there were more proteins identified with proteome-wide significance in the 3D-CLUMP results (**Figure 5**). The number of proteins identified uniquely as proteome-wide significant in the 3D test was between 3 times to 10.5 times more than the number uniquely identified in the 1D test. In some instances, the same protein was identified in both the 1D and 3D tests (e.g., PIK3CA in TCGA BRCA, ALG13 in TCGA COAD, PACS1 in TCGA COAD). However, most of the time the proteins discovered were unique to the specific test showing a benefit to using 3D structures in assessment of clustering of missense variation.

**Figure 5:**
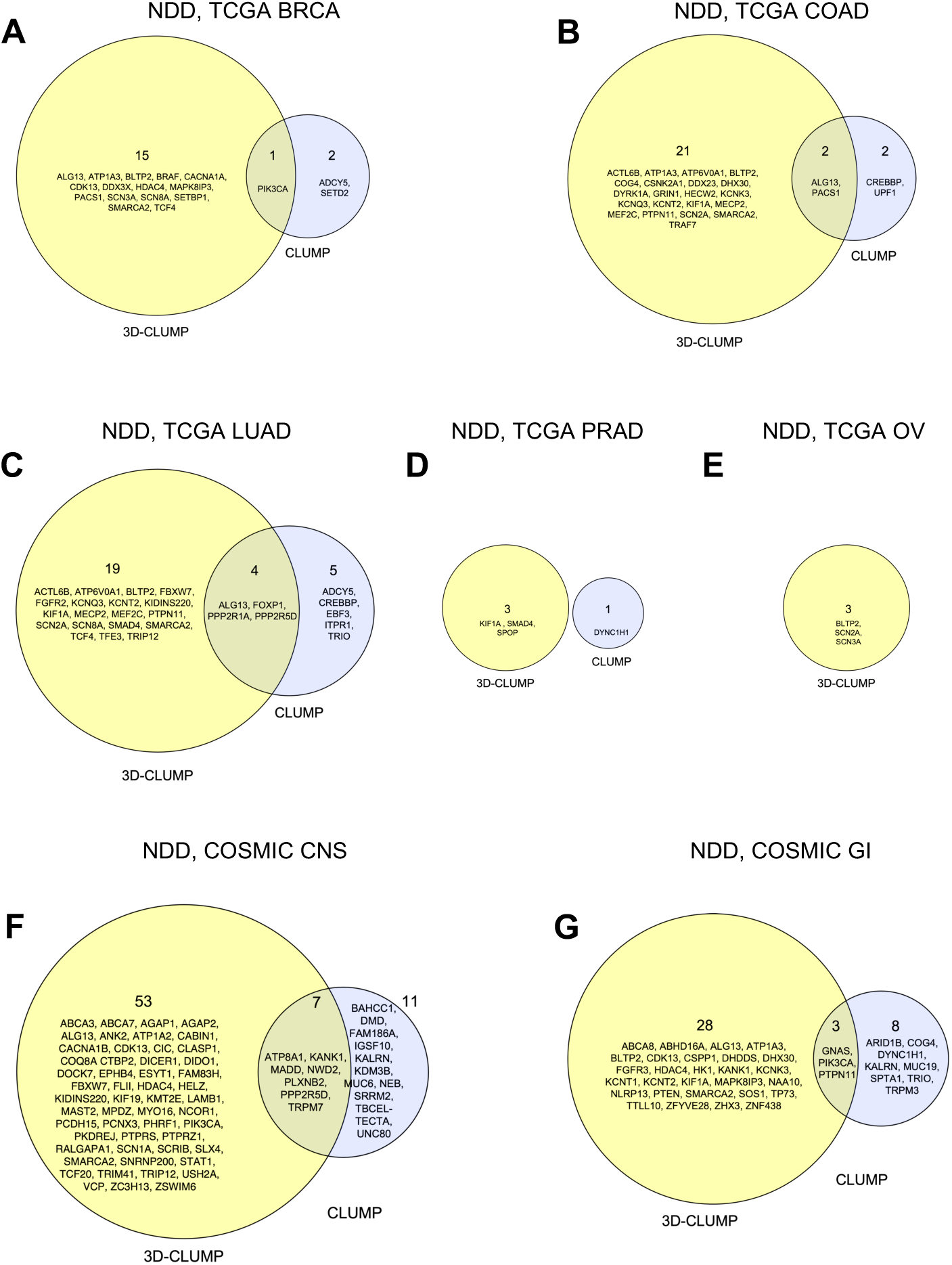
Discovery is Greater with 3D Structures. Shown are proteins exhibiting proteome-wide significance for clustering in either NDDs or the specified cancer type using the 3D-CLUMP and/or CLUMP methods.

### Protein-Protein Interaction Enrichment of Proteins with Significant Missense Clustering

For the proteins enriched for clustering in NDDs and cancer we tested for enrichment of protein-protein interactions (PPIs) in STRING-DB (**Figure 6**). For the proteins with enrichment in NDDs, we identified significant PPIs (number of nodes = 57, number of edges = 123, expected number of edges = 41, p < 1 x 10^−16^) with enrichment of proteins involved in the BAF complex and proteins that function as channels. In the proteins identified in the cancer analysis, we also so an enrichment of PPIs (number of nodes = 78, number of edges = 51, expected number of edges = 33, p = 2.5 x 10^−3^) and enrichment of proteins involved in ATP-related activities.

**Figure 6:**
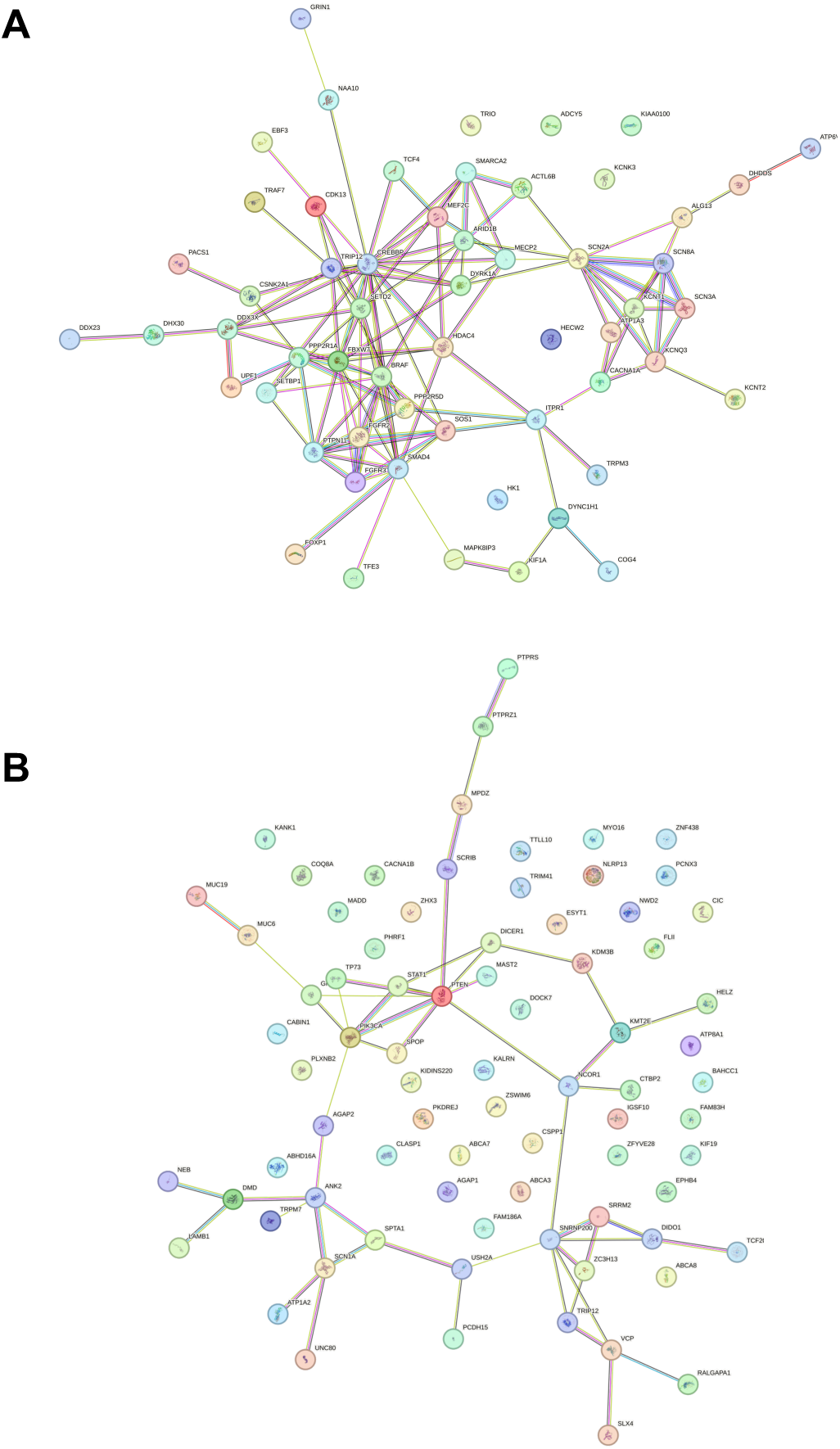
Protein-Protein Interaction Enrichment of Proteins with Significant Missense Clustering. A) PPI network of proteins with proteome-wide significant clustering in NDDs (number of nodes = 57, number of edges = 123, expected number of edges = 41, p < 1 x 10^−16^). B) PPI network of proteins with proteome-wide significant clustering in cancer (number of nodes = 78, number of edges = 51, expected number of edges = 33, p = 2.5 x 10^−3^).

### Comparison to Known NDD Genes

There are 379 NDD genes known to be genome-wide significant for enrichment of DNVs (36, 43). We examined what the status of these genes were in this study (**Figure 7**). There were 50 genes that were known to be significant in NDDs and exhibited significant missense DNV clustering in NDDs in this study. There were 13 genes that were known to be significant in NDDs and exhibited significant missense DNV clustering in cancer in this study. One gene (*TRIP12*) was found in both of these results. TRIP12 was significant in NDDs in the comparison to LUAD and TRIP12 is significant in COSMIC CNS in comparison to NDDs. This results in 62 genes exhibiting clustering in the known NDD set. The remaining 319 proteins that were not found significant by CLUMP or 3D-CLUMP either had a small number of independent missense variants (<6) for CLUMP/3D-CLUMP testing, or were too large to build an AlphaFold structure with our compute memory limitation of 700 GB. There were 135 proteins with <6 independent missense variants and 25 for which we could not generate AlphaFold structures. There were 157 proteins that were not significant in any of our tests and likely did not harbor any clustered missense variation.

**Figure 7:**
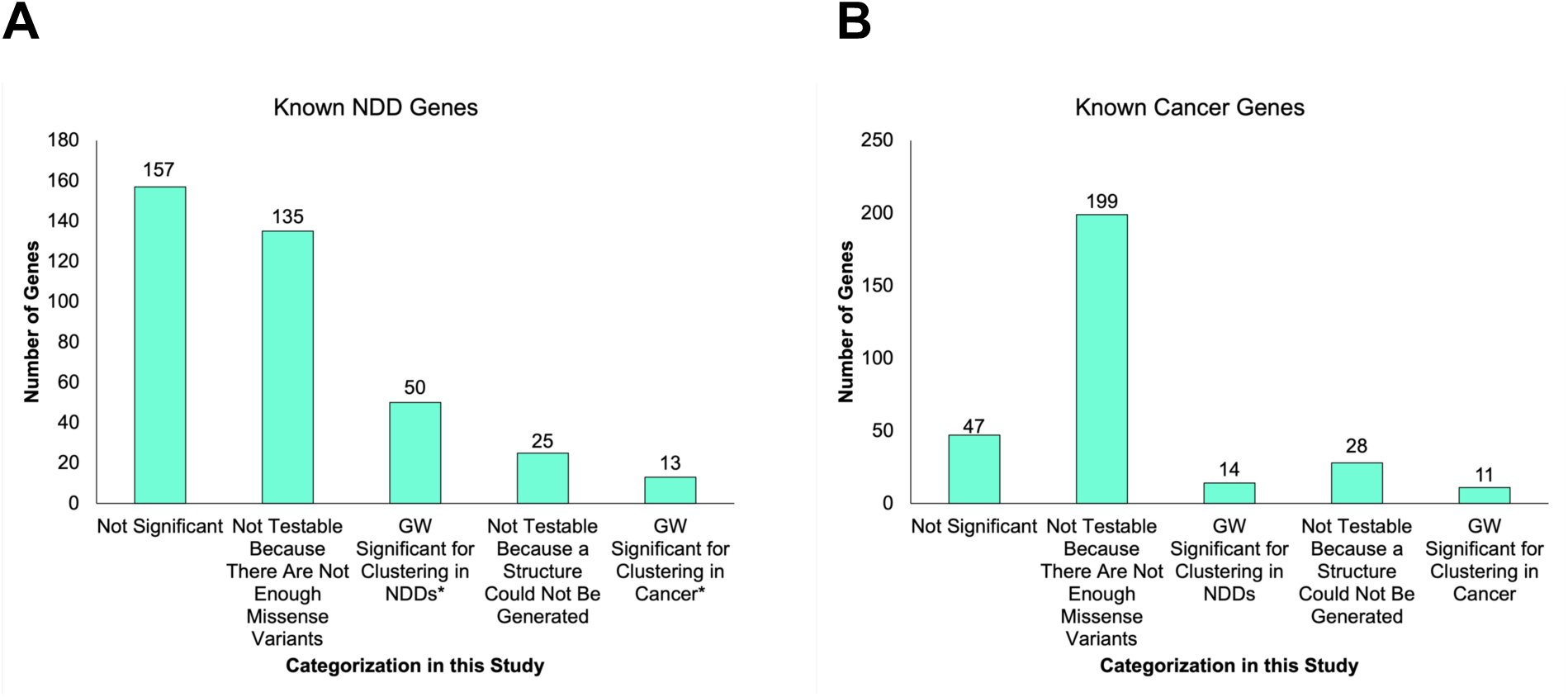
Comparison to Known NDD and Known Cancer Genes. A) Shown is the distribution of protein results in our study and comparison to 379 known NDD genes. **\***TRIP12 is significant in NDDs in comparison to LUAD and is significant in COSMIC CNS comparison to NDDs. B) Shown is the distribution of protein results in our study and comparison to 299 known Cancer genes

Based on the analyses above, for the 379 known NDD proteins there were 62 (16.4%) with clustering based on CLUMP/3D-CLUMP analyses, and 160 (42.2%) were not testable either because they did not have enough missense variants to perform the analyses or a protein structure could not be made, and 157 (41.4%) were not significant for clustering by any test.

### BLTP2 as a Novel Proteome-Wide Significant Protein in NDDs

There were seven proteins (ACTL6B, BLTP2, DHX30, KCNT2, MAPK8IP3, SCN3A, SOS1) with significant clustering in NDDs that were not previously genome-wide significant for DNVs in NDDs. Six (ACTL6B, DHX30, KCNT2, MAPK8IP3, SCN3A, SOS1) of these proteins have been implicated in rare forms of NDDs. ACTL6B is involved in in “developmental and epileptic encephalopathy 76” (OMIM #618468) and “intellectual developmental disorder with severe speech and ambulation defects” (OMIM #618470). DHX30 is involved in “neurodevelopmental disorder with variable motor and speech impairment” (OMIM #617804). KCNT2 is involved in “developmental and epileptic encephalopathy 57” (OMIM #617771). MAPK8IP3 is involved in “neurodevelopmental disorder with or without variable brain abnormalities” (OMIM #618443). SCN3A is involved in “developmental and epileptic encephalopathy 62” (OMIM #617938) and “epilepsy, familial focal, with variable foci 4” OMIM #617935). SOS1 is involved in “Noonan syndrome 4” (OMIM #610733).

One protein (BLTP2) has not been implicated in NDDs before (**Figure 8**). BLTP2 was previously known as KIAA0100. We identified significant clustering of missense variants in NDDs in this protein using our 3D-CLUMP tool. It was significant, with a 3D-CLUMP Score in NDDs 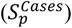 of 0.28, in the comparison to BRCA (p = 1 × 10^−7^), COAD (p = 2 × 10^−7^), LUAD (p = 5 × 10^−7^), OV (p = 1.2 × 10^−6^), and COSMIC GI (p < 1 × 10^−7^). In particular, this protein had seven amino acid changes on isoform NP_001350756 with three at amino acid position 1487 and one each at amino acid positions 605, 705, 1253, and 1483. Running a conserved domain prediction on the protein, we found that the variants at positions 1483 and 1487 are within the Apt1 domain of this protein that is predicted to be involved in localization of the protein to the Golgi body. Since there were three individuals with a missense variant at amino acid position 1487, we also checked the gnomAD database for this variant (17-28619915-C-T). This allele is seen in 3 individuals in our study (3 alleles / 79766 total alleles, allele frequency = 3.76 × 10^−5^) and is seen in 1 individual assessed in gnomAD (1 allele / 780820 total alleles in genome+exome samples, allele frequency = 1.28 × 10^−6^). This allele is enriched in the NDD cohort (Fisher’s Exact Test p = 2.96 × 10^−3^, OR = 29.4). To further examine missense variants at this position, we examined another publication consisting of an independent set of 13,189 individuals with DNVs from whole-exome sequencing data from the SPARK autism cohort (42). In this cohort, there was one individual with a missense variant at the same amino acid position (17-28619916-G-A) resulting in an Arginine to Tryptophan change (1 allele / 26378 alleles, allele frequency = 3.79 × 10^−5^) and this same variant is seen in 3 individuals in gnomAD (3 allele of 1613876 total alleles, allele frequency = 2.48 × 10^−6^) in genome+exome. This allele is also enriched in NDDs (Fisher’s Exact Test p = 8.30 × 10^−4^, OR = 15.2). In addition to the enrichment in NDDs, it is relevant to note that the Arginine is also highly conserved in several species (**Figure 8**).

**Figure 8:**
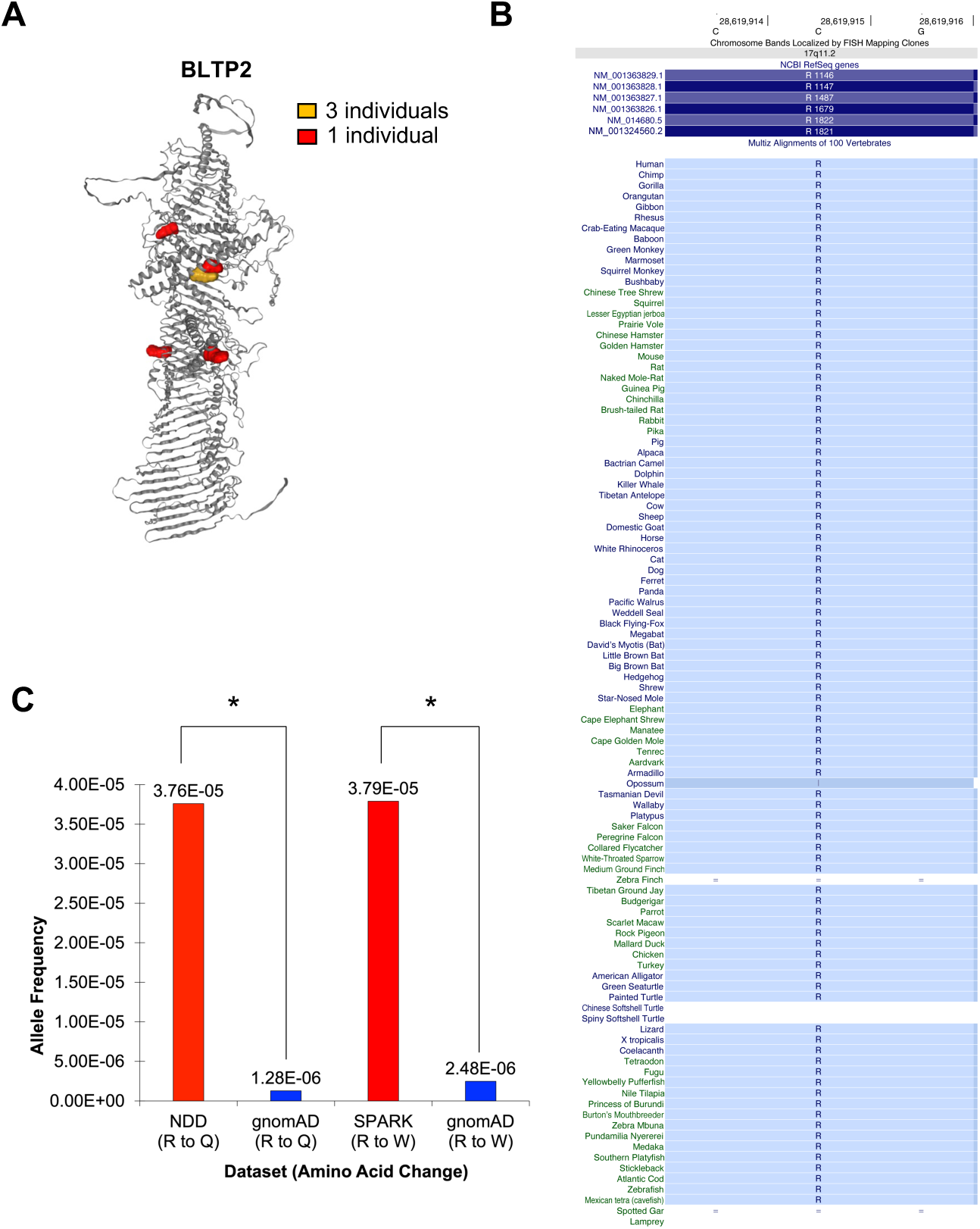
Discovery of BLTP2. A) Shown are missense DNVs observed in individuals with NDDs on the BLTP2 protein structure (NP_001350756.1). This protein had seven amino acid changes with three at amino acid position 1487 and one each at amino acid positions 605, 705, 1253, and 1483. B) The Arginine at position 1487 is highly conserved across several species. C) The Arginine to Glutamine missense variant is significantly enriched in NDDs (Fisher’s Exact Test p = 2.96 × 10^−3^, OR = 29.4). Another missense variant (Arginine to Tryptophan) was identified at this amino acid position in an independent NDD cohort (SPARK) and is also enriched in NDDs (Fisher’s Exact Test p = 8.30 × 10^−4^, OR = 15.2)

There are few publications characterizing BLTP2 (KIAA0100) (61–71). None of these papers implicate BLTP2 in NDDs. However, what is known is that it is a member of the Bridge-Like Lipid Transport Protein family. These proteins are important for transfer of lipids and other members of the protein family have been implicated in neurodevelopmental and neurodegenerative disorders (72, 73). A recent preprint has also indicated a role for BLTP2 in the regulation of primary cilia (74); an area of molecular interest in neurodevelopmental disorders (75).

### Comparison to Known Cancer Driver Genes

Estimates of the number of mutation-based cancer driver genes have varied over the years, but a commonly used source is the paper from the Cancer Genome Atlas (TCGA) (49). This study identified 299 driver genes, of which 259 were the result of consensus predictions of 26 computational tools. A limited number of these genes met the criteria for analysis in our study, given our threshold for independent missense mutations and limitations of our AlphaFold modeling described above (**Figure 7**). Of these genes, 14 were significantly clustered in NDDs and 11 were significantly clustered in cancers. There were 199 proteins that were not testable because there were not enough missense variants to run the test and 28 could not be tested because a structure could not be generated for the protein. There were 47 proteins that were tested and not significant.

Based on the analyses above, for the 299 known cancer proteins there were 25 (8.4%) with clustering based on CLUMP/3D-CLUMP analyses, and 227 (75.9%) were not testable either because they did not have enough missense variants to perform the analyses or a protein structure could not be made, and 47 (15.7%) were not significant for clustering by any test.

## 62 potential proteome-wide significant genes in cancers not discovered by TCGA

Of the remaining genes, eleven in the Bailey list and 68 not in the list were found to have proteome-wide significant differential clustering in one of our cohorts (COSMIC-CNS, COSMIC-GI, TCGA-LUAD, TCGA-BRCA, or TCGA-PRAD) when compared to NDD variants based on CLUMP/3D-CLUMP analyses. Six of the 68 genes not on the Bailey list had been proposed as a driver in the literature, with an additional 11 proposed as prognostic or as a therapeutic target in a particular cancer type. Two genes were paralogs of drivers on the Bailey list.

### Proteins Requiring Careful Consideration for Prognostics and Therapeutics

There were 220 known NDD proteins that we could test for missense clustering in NDDs or cancer (see above, **Figure 7**). Of these, 50 were proteome-wide significant for clustering in NDDs. However, there were 13 (5.9%) that were proteome-wide significant for clustering in cancer (AGAP2, ANK2, CLASP1, GNAS, KIDINS220, KMT2E, PIK3CA, PTEN, SCN1A, SRRM2, TCF20, TRIP12, UNC80). These proteins will need to be specially considered when thinking about functional, prognostic, and therapeutic aspects of the variants within them in different phenotypes. Likewise, there were 72 known cancer proteins that we could test for missense clustering in NDDs or cancer (see above, **Figure 7**). Of these, 11 were proteome-wide significant for clustering in cancer. However, there were 14 (19.4%) that were proteome-wide significant for clustering in NDDs (BRAF, CACNA1A, CREBBP, DDX3X, FBXW7, FGFR2, FGFR3, KIF1A, PPP2R1A, PTPN11, SETBP1, SETD2, SMAD4, SOS1). These proteins will also need to be specially considered when thinking about functional, prognostic, and therapeutic aspects of the variants within them in different phenotypes.

## DISCUSSION

An outstanding question in the genomics of NDDs is why are several genes identified in NDDs also identified in cancer? In particular, genes identified in both are involved in molecular processes including chromatin remodeling and transcription (58). Other interesting observations include microcephaly and macrocephaly as a result of variation in some genes in NDDs. This has been compared to the cellular proliferation and differentiation related processes in cancer. Another area of interest has been in genome maintenance (57). Several hypotheses have been put forward for the overlap in genes (58).

In this study, we explore the hypothesis that genes with missense variants in NDDs and cancer have a different variant pattern in NDDs and in cancer. Three main options were considered at the protein-level including clustering of missense variants in NDDs and not in cancer, clustering of missense variants in cancer and not in NDDs, and no clustering of missense variants in NDDs or cancer. Since we focused on clustering of missense variation at the level of each protein, we utilized two strategies including examination of clustering on the 1D protein structure and clustering on the 3D protein structure. Our existing method CLUMP (52) was utilized to look for proteome-wide significance of clustering in 1D in a case-control design. We also developed the computational tool 3D-CLUMP that can perform proteome-wide significant case-control analysis in three-dimensional protein structure space. This computational tool is open-source, available on GitHub (https://github.com/TNTurnerLab/3D-CLUMP), and will be beneficial to others wanting to test for clustering of variants on 3D protein structures in these and other phenotypes. As part of this work, we also generated AlphaFold structures for >4000 proteins with enough missense variation to be tested in our study. These structures were deposited in ModelArchive (https://www.modelarchive.org/doi/10.5452/ma-tur-clump) and will also be beneficial as a resource to the research community. We showed in this study in the comparison of CLUMP (1D) and 3D-CLUMP that using 3D structures boosts our power for discovery of proteins with clustering of missense variation.

Several outcomes of our study are novel and intriguing with regard to missense variants in NDDs and cancer. By comparing missense variants in NDDs to those in cancer, we identified proteins where there was significant clustering of missense variants in NDDs, proteins where there was significant clustering of missense variants in cancer, and proteins with no clustering in NDDs or cancer. This is an important discovery because it points to specific proteins where there are differences in the patterns of missense variation in the two phenotypes. This is another important resource for researchers studying the two phenotypes and will provide important information relevant in functional assessment of variation and in prognostics. While many of the genes we identified in NDDs have been identified in more broad searches looking at DNV enrichment of likely-gene disrupting variation and missense variation irrespective of clustering (36, 43), we did also identify one new proteome-wide significant protein (BLTP2) for NDDs. Overall, our work provides novel insights into patterns of missense variation in NDDs and cancer.

There are a few caveats to our study. One caveat is that there are some proteins where we do not have enough missense variants to perform the statistical test, and this is something that will be approachable with increased sample sizes in both NDDs and cancer. Another caveat is regarding the protein structures themselves. For some proteins, the protein isoform may not be supported by experimental evidence, the structure could not be made, or for some a high confidence structure could not be made. Finally, our statistical test does not currently identify when the proteins have clustered missense variants in both NDDs and cancer but the highly clustered regions are different in the two phenotypes. Potential future work would address each of these caveats to provide further insights into this question. One other interesting future direction is to explore the clustering of missense variants with regard to PPIs. We showed an enrichment of these and careful consideration of clusters at the interfaces of these PPIs would be useful.

## Data Availability

All data produced in the present work are contained in the manuscript

https://www.modelarchive.org/doi/10.5452/ma-tur-clump

https://github.com/KarchinLab/CLUMP

https://github.com/TNTurnerLab/3D-CLUMP

https://github.com/TNTurnerLab/clustering_in_cancer_vs_ndd_paper

## ACKNOWLEDGMENTS

This work was supported by grants from the National Institutes of Health (R01MH126933 to T.N.T., R00MH117165 to T.N.T., P50HD103525 to T.N.T. as a Member and Scientific Liaison in the Washington University in St. Louis Intellectual and Developmental Disabilities Research Center), the ITCR program at the National Cancer Institute (U24CA258393 to R.K.), the Simons Foundation (Award #734069 to T.N.T.), and funds from the Washington University in St. Louis McDonnell Center for Cellular and Molecular Neurobiology to T.N.T. Thank you also to Dan Western for his participation on the project during his rotation in the Turner Laboratory.

## CRedIT AUTHOR STATEMENT REGARDING AUTHOR CONTRIBUTIONS

**Jeffrey K. Ng:** Methodology, Software, Formal analysis, Investigation, Data Curation, Writing - Original Draft, Writing - Review & Editing, Visualization. **Yilin Chen:** Methodology, Software, Formal analysis, Investigation, Data Curation, Writing - Original Draft, Writing - Review & Editing, Visualization. **Titilope M. Akinwe:** Methodology, Formal analysis, Investigation, Data Curation, Writing - Original Draft, Writing - Review & Editing, Visualization. **Hillary B. Heins:** Formal analysis, Writing - Review & Editing. **Elvisa Mehinovic:** Formal analysis, Writing - Review & Editing. **Yoonhoo Chang:** Formal analysis, Writing - Review & Editing. **Zachary Payne:** Formal analysis, Writing - Review & Editing. **Juana G. Manuel:** Visualization, Writing - Review & Editing. **Rachel Karchin:** Methodology, Software, Formal analysis, Investigation, Resources, Data Curation, Writing - Original Draft, Writing - Review & Editing, Visualization, Supervision, Project administration, Funding acquisition. **Tychele N. Turner:** Conceptualization, Methodology, Software, Formal analysis, Investigation, Resources, Data Curation, Writing - Original Draft, Writing - Review & Editing, Visualization, Supervision, Project administration, Funding acquisition.

## DATA AND SOFTWARE AVAILABILITY STATEMENT

The AlphaFold structures for this project have been deposited in ModelArchive under accession number “ma-tur-clump” at https://www.modelarchive.org/doi/10.5452/ma-tur-clump. The code for this paper is available at:

**CLUMP:** https://github.com/KarchinLab/CLUMP

**3D-CLUMP:** https://github.com/TNTurnerLab/3D-CLUMP

**AlphaFold Structure and 3D Protein Plot Generation:** https://github.com/TNTurnerLab/clustering_in_cancer_vs_ndd_paper

